# The Genetic Landscape and Epidemiological Characteristics of Inherited Retinal Diseases in the Chinese Population

**DOI:** 10.64898/2026.05.27.26354224

**Authors:** Bing Zeng, Zhonghui Cui, Siting Zhou, Weiwei Dai

## Abstract

**Background:** Inherited Retinal Diseases (IRDs) are a group of genetically heterogeneous blinding conditions. Major global genomic reference databases are disproportionately enriched for individuals of European ancestry. This underrepresentation creates a significant bias that impedes the accuracy of genetic diagnosis in the Chinese population. This study aims to address this limitation by constructing a comprehensive genetic landscape of IRDs using large-scale deep-sequencing data from a large Chinese cohort.

**Methods:** The study leveraged variant data primarily from 10,588 individuals in the China Metabolic Analytics Project (ChinaMAP) and cross-referenced findings against multiple national and international databases. We systematically curated variants within a targeted panel of 291 IRD-associated genes. Variant pathogenicity was assessed using a comprehensive pipeline integrating InterVar-automated classification based on 2015 American College of Medical Genetics and Genomics/Association for Molecular Pathology (ACMG/AMP) guidelines, ClinVar evidence (review status ≥ 1 star), and manual literature curation. We delineated the mutational spectrum, identified population-enriched pathogenic/likely pathogenic (P/LP) variants, and analyzed the distribution characteristics of IRD-associated highly-mutated genes. Furthermore, we calculated the carrier frequencies (CF) and genetic prevalence (GP) of autosomal recessive(AR)-IRD genes in the Chinese population.

**Results:** The study revealed a highly concentrated genetic landscape for AR-IRDs in the Chinese population, with *ABCA4* and *USH2A* emerging as the primary drivers of the genetic burden. This finding aligns with previous Chinese cohorts but contrasts with global databases, where genes such as the X-linked *RPGR* are more prevalent. In contrast, autosomal dominant (AD)-IRDs exhibited high locus heterogeneity, with pathogenic variants dispersed across numerous genes (e.g., *COL2A1* and *MFN2*). We identified a series of P/LP variants that were either high-frequency or significantly enriched in the Chinese population, such as *CNGB1* (p.P530R) and specific recurrent alleles in *ABCA4* and *CYP4V2*. The estimated cumulative CF for AR-IRDs was 1 in 5.60, and the theoretical total GP was 1 in 2,624.67, based on the ChinaMAP data.

**Conclusion:** By integrating the ChinaMAP dataset with diverse genomic resources, this study provides a genetic landscape of IRDs in the Chinese population. Our analysis shows a concentrated mutational spectrum in AR-IRDs, contrasting with the pronounced heterogeneity in AD-IRDs. These findings, including population-specific pathogenic variants and refined prevalence estimates, provide a resource for precision diagnostics, genetic counseling, expanded carrier screening (ECS), and public health policy development in China.

## 1. Introduction

Inherited retinal diseases (IRDs) are a group of clinically and genetically heterogeneous Mendelian disorders encompassing various phenotypes^[1]^, including retinitis pigmentosa^[2]^, Leber congenital amaurosis^[3]^, and Stargardt disease^[4]^. They are caused by pathogenic mutations in at least 277 nuclear and mitochondrial genes^[5]^, which trigger the progressive degeneration of different retinal regions, ultimately leading to severe vision loss or irreversible blindness^[1]^. Given their genetic basis, a precise genetic diagnosis is important for elucidating etiology, predicting prognosis, guiding genetic counseling, and developing targeted interventions such as gene therapy^[5,6]^.

Next-generation sequencing has accelerated the discovery of IRD-causative variants^[7,8]^, but the interpretation of these variants depends heavily on reference population databases. Large-scale reference databases remain predominantly of European origin^[9]^, which limits their applicability to non-European populations. This data bias leads to discordant variant interpretation when applied to the Chinese population, a major component of the East Asian ancestry group^[8]^. Pathogenic variants that are common or enriched in Chinese individuals are often rare or absent in European reference cohorts, directly compromising diagnostic accuracy^[10]^. Previous studies have identified IRD-related variants in Chinese patient cohorts, but most have been based on relatively small clinical samples^[11,12]^ , and their findings may not fully represent the genetic landscape of the general population. As a result, variants unique to or enriched in the Chinese population, such as those in *USH2A* and *EYS*^[8,13]^, may be under-appreciated.

To address this gap, we leveraged high-depth (40X) whole-genome sequencing (WGS) data from 10,588 individuals in the ChinaMAP^[14]^, integrated with multiple national and international databases, to characterize a comprehensive genetic landscape of IRDs in the Chinese population to date. Specifically, we aimed to: (1) delineate the mutational spectrum of IRDs, contrasting the genetic landscape of autosomal recessive (AR) forms with autosomal dominant (AD) forms; (2) identify pathogenic or likely pathogenic (P/LP) variants that are enriched in the Chinese population; and (3) estimate carrier frequencies (CF) and genetic prevalence (GP) for AR-IRDs based on population-level data.

## 2. Methods

The overall analytical workflow is summarized in Figure 1. Briefly, we first identified P/LP variants in IRD-associated genes using the ChinaMAP cohort as the primary dataset. Pathogenicity was assessed through an integrated pipeline combining InterVar-based ACMG/AMP classification, ClinVar evidence, and manual literature curation. We then cross-validated the identified variants across multiple Chinese and global population databases to characterize the mutational spectrum, identify population-enriched variants, and estimate CF and GP for autosomal recessive IRDs.

**Figure 1.**
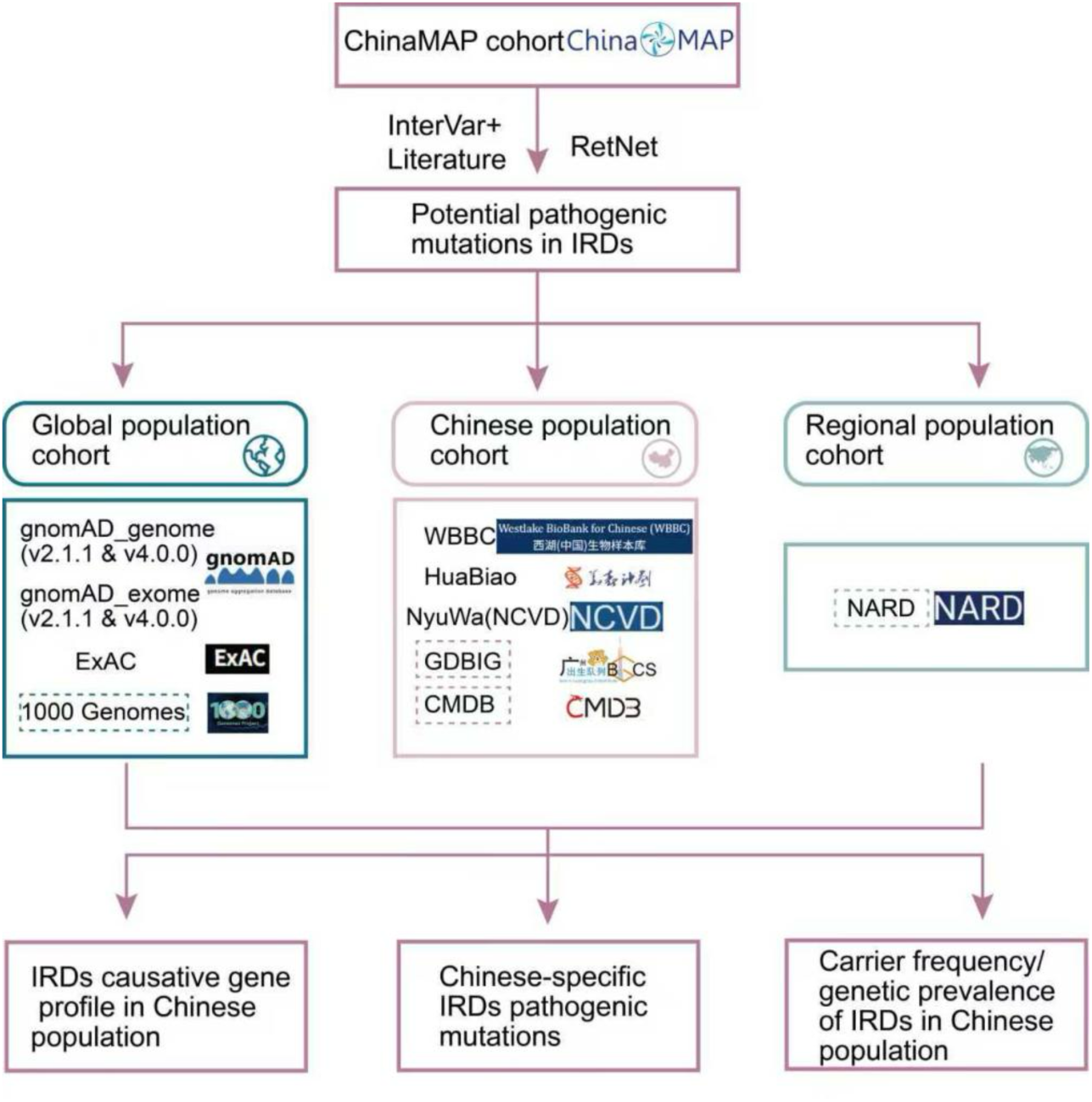
The analysis flowchart of this study. Solid-line boxes indicate cohorts used for variant identification and allele frequency calculation; dashed-line boxes indicate cohorts used only for retrieving allele frequencies.

### 2.1 Data Sources and Cohorts

Demographic and variant data for the Chinese population were primarily derived from the ChinaMAP database (http://www.mbiobank.com/; version: 2020-03.beta), which represents the largest publicly available high-depth whole-genome sequencing cohort in China. To ensure the robustness of our findings, we performed cross-cohort validation and cross-population comparisons using several additional datasets. These included other large-scale Chinese cohorts (Westlake BioBank for Chinese population [WBBC], “HuaBiao” Project [HuaBiao], NyuWa Chinese Population Variant Database [NCVD], Genome Database of Born In Guangzhou Cohort study [GDBIG], China Millionome Database [CMDB]) and major global population databases (Genome Aggregation Database [gnomAD] exome and genome V2.1.1 & V4.0.0, Exome Aggregation Consortium [ExAC], The 1000 Genomes Project [1000G], and Northeast Asian Reference Database [NARD]). Detailed descriptions of all cohorts are provided in Supplementary Table 1. Given its substantial sample size and high-depth sequencing, the ChinaMAP cohort served as the primary dataset for all main analyses. All genomic analyses were standardized to the hg38 reference genome; variant data from cohorts based on the hg19 version were lifted over by the CrossMap package (V0.7.0) when necessary.Principal Component Analysis (PCA) demonstrated a clear genetic stratification among the analyzed databases (Supplementary Figure S1).

### 2.2 Selection and Annotation of IRD-Associated Genes

A comprehensive list of 291 genes associated with IRDs was compiled from the RetNet database (V2022107; https://web.sph.uth.edu/RetNet/). The inheritance patterns for these genes were annotated based on the Online Mendelian Inheritance in Man (OMIM) database (https://www.omim.org/) and classified into seven modes: AR, AD, X-linked recessive (XLR), X-linked dominant (XLD), digenic dominant, mitochondrial, and unknown. For genes with multiple inheritance annotations (e.g., both AR and AD), we implemented a hierarchical prioritization strategy favoring recessive classifications (AR over AD; XLR over XLD). This conservative approach ensured the biological validity of CF and GP estimations, as inclusion of dominant genes would confound the differentiation of asymptomatic carriers. Genes lacking a documented inheritance mode in OMIM were classified as ’unknown’ (Supplementary Table 2). These, alongside genes with exclusively dominant or X-linked inheritance, were excluded to establish a finalized panel of 209 high-confidence AR-IRD genes.

### 2.3 Pathogenicity Assessment of Sequence Variants

The pathogenicity of IRD-associated variants was assessed with InterVar (V1.0.8), which programmatically implements the 2015 American College of Medical Genetics and Genomics/Association for Molecular Pathology guidelines (ACMG/AMP)^[15–18]^. Variants classified as ’Pathogenic’ or ’Likely Pathogenic’ (P/LP) by InterVar constituted our primary analysis set (Set 1). To enhance comprehensive coverage, this set was augmented with a curated list of established IRD-associated P/LP variants derived from the ClinVar database (review status ≥ 1 star) and a comprehensive literature review^[1]^. All external variants were lifted over to the GRCh38 (hg38) assembly to ensure coordinate consistency. This merged list formed the final comprehensive pathogenic variant set used for all subsequent analyses.

To delineate the distinct genetic architecture of IRDs in the Chinese population, we implemented a dual-threshold filter to identify population-specific enriched variants. A variant was defined as “Chinese-population-specific” based on the following criteria: (1) an allele frequency (AF) greater than 0.001 in at least one of the analyzed Chinese population databases (e.g., ChinaMAP, WBBC, or HuaBiao); and (2) an AF less than 0.0001 in all non-Chinese global reference databases (excluding the East Asian subpopulation data where applicable). This filtering strategy effectively differentiated founder effects or localized mutational burdens from rare sporadic mutations or globally prevalent polymorphisms.

### 2.4 Statistical Analysis and Visualization

A series of analytical approaches was employed to characterize the genetic architecture of IRDs across populations. First, Bump Charts were used to visualize the shifting rank order of the top 21 genes (union of top 10 per database), thereby revealing cross-population trends in gene-level mutation burden.

To quantify the concentration of pathogenic drivers, we performed Lorenz curve analysis and calculated the Gini coefficient (G), where values approaching 1 indicate a highly concentrated mutational burden. Databases with sparse detection of AD-associated variants (total count < 10) were excluded to ensure data fidelity. All analyses were implemented using the ineq and ggplot2packages in R. Applying this filter, the NARD, WBBC, and HuaBiao databases were excluded from the AD-IRD Lorenz curve and Gini coefficient analyses due to insufficient distinct P/LP variants (each < 10).

Given the variable sequencing depths across cohorts, relative allelic diversity (δ) was calculated for each gene. The stability of the mutational spectrum was then assessed using the coefficient of variation (CV). To avoid inflation of CV from undetected variants, any gene showing zero P/LP variants across all databases within a given cohort scale (e.g., all Chinese cohorts) was excluded from CV calculation and permutation analysis for that scale.

Permutation tests (1,000 iterations) were conducted to determine whether the observed CV reflected biological constraints rather than random noise. A null distribution of mean CVs was generated by randomly shuffling gene–inheritance labels across the dataset, and empirical P-values were derived from the position of the observed mean CV relative to this stochastic distribution. This approach specifically aimed to unmask the “zero-inflation” effect in smaller datasets. Tests were stratified by three cohort scales: (1) all seven databases, (2) Chinese cohorts only (ChinaMAP, HuaBiao, WBBC), and (3) global cohorts only (ExAC, gnomAD V2.1.1 and V4.0.0). This stratification allowed us to evaluate whether the mutational stability of AR and AD genes is influenced by population composition or database scale.

To explore the structural impact of population-specific variants, we mapped P/LP mutations onto linear protein structures using Lollipop plots. Functional domain annotations were retrieved from UniProt and InterPro. Variant frequencies were plotted on the Y-axis (with a split-scale for high-frequency alleles) to identify mutational hotspots within key biochemical motifs.

Finally, to facilitate cross-database comparisons that involved varying sample sizes and sequencing depths, we calculated relative allelic diversity for each gene as the number of distinct P/LP variants in that gene divided by the total number of P/LP variants in the database. Row-wise Z-score normalization was applied to the resulting matrix, followed by hierarchical clustering using Euclidean distance and complete linkage. Heatmaps were generated with the pheatmap package in R (version 1.0.12).

### 2.5 Calculation of CF and GP

Following the pathogenicity assessment pipeline (Section 2.3), a total of 830 unique P/LP variants across 209 AR-IRD genes were identified in the ChinaMAP cohort, forming the basis for all CF and GP calculations. An analogous procedure applied to the WBBC cohort yielded 552 unique P/LP variants. Both CF and GP for AR-IRDs were derived from cumulative allele frequencies of these P/LP variants.

For an individual pathogenic variant *i* with an allele frequency *AFᵢ*, the variant-level CF (*CF_variant, i_*) was calculated as:

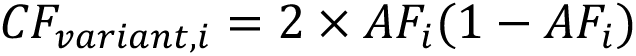

For a gene *i* harboring *n* distinct pathogenic variants, the total gene-level CF (*CF_gene, i_*) was calculated as:

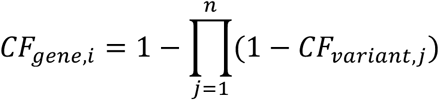

Because individual-level phasing data and explicit zygosity information are unavailable in ChinaMAP and WBBC, we adopted a probabilistic matrix model based on AF to estimate the GP for each AR-IRD gene. Under the assumptions of random mating and independence among pathogenic variants, GP was calculated by aggregating probabilities for both homozygous and compound heterozygous states.

Specifically,we employed a product-based algorithmic model that leverages allele matrices derived from the ChinaMAP dataset. For any two pathogenic variants *i* and *j* within the same gene, the probability of an individual harboring both (thereby manifesting the disease) was derived from the product of their respective CF (*CF_i_* and *CF_j_*), scaled by the Mendelian segregation ratio of 1/4. The gene-specific GP (*GP_gene_*) is therefore approximated by:

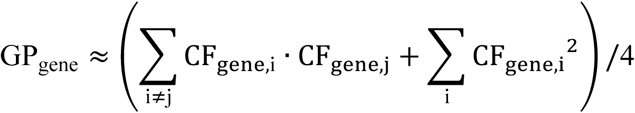

To obtain the cumulative CF (*CF_cumulative_*) for the entire panel of *m* = 209 AR-IRD genes without overcounting, we applied the principle of independent probabilities:

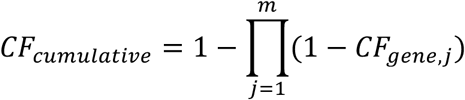

Finally, the total GP (*GP_total_*) for the entire AR-IRD disease group was estimated by summing the individual gene-level GPs:

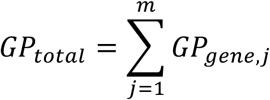

This additive method assumes locus heterogeneity, where mutations in different genes cause clinically overlapping but genetically distinct disorders, thereby representing the overall mutational burden of IRDs^[19,20]^. Given the rarity of IRD-causative variants, linkage disequilibrium between distinct pathogenic alleles within the same gene was assumed to be negligible.

## 3. Results

### 3.1 Causative Gene Profile of IRD in the Chinese Population

Analysis of P/LP variants across 291 IRD-associated genes in the ChinaMAP cohort revealed a highly concentrated mutational landscape. *ABCA4* was the principal contributor, harboring 67 distinct P/LP variants, followed by *USH2A* (n=55), *CRB1* (n=22), *EYS* (n=22), and *CNGA3* (n=21) (Table 1). Hierarchical clustering of the top 20 genes separated Chinese cohorts (ChinaMAP, HuaBiao, WBBC) from predominantly European cohorts (gnomAD, ExAC) (Figure 2A). After normalizing for database size, *CYP4V2* and *CNGB1* were markedly enriched in Chinese datasets but depleted in predominantly European databases (Figure 2B).

**Table 1.**
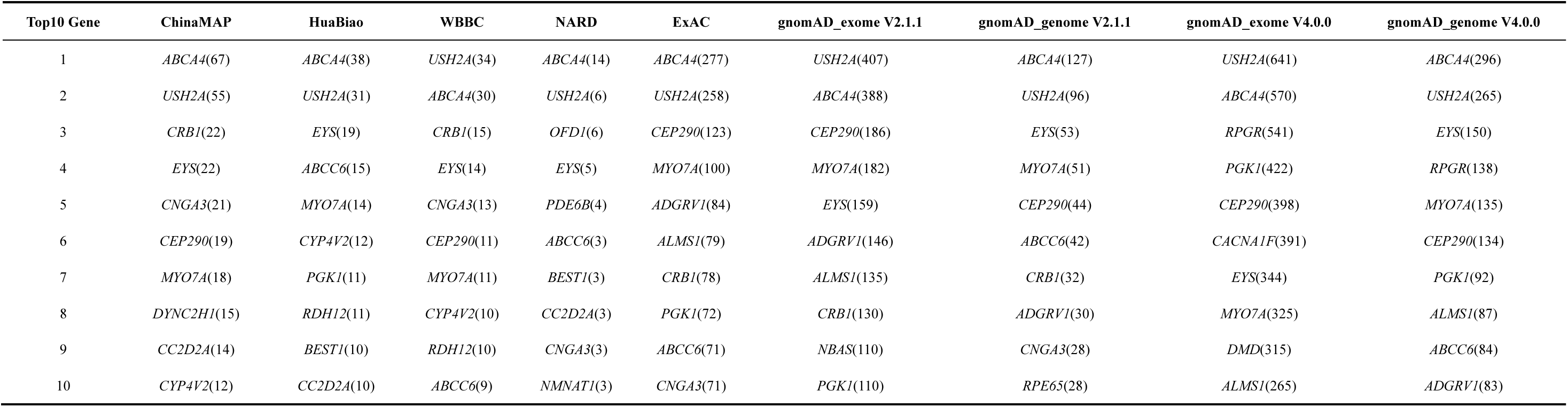
Comparative profile of the top 10 IRD-associated genes harboring P/LP variants.

**Figure 2.**
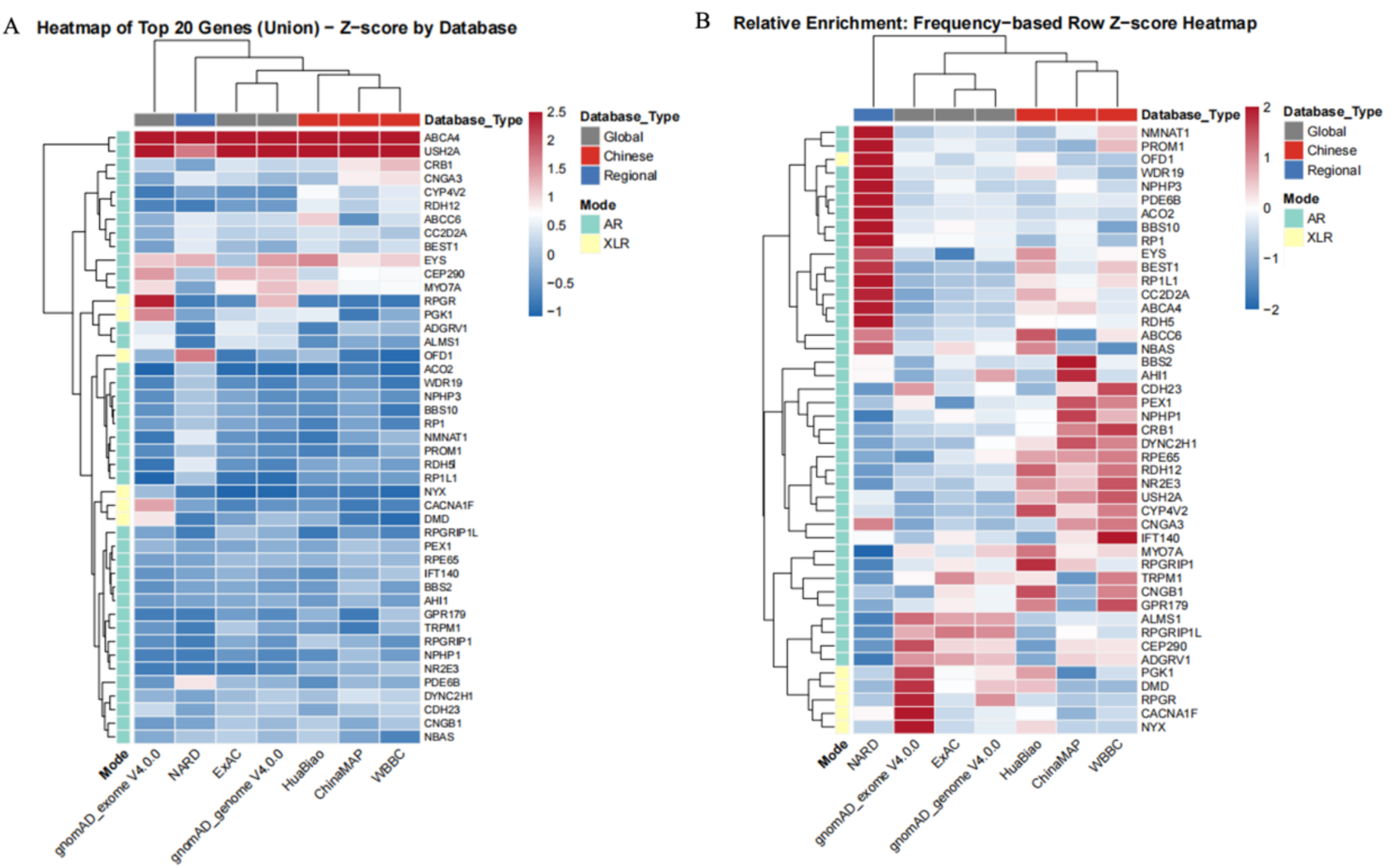
Landscape of P/LP variants in top 20 IRD genes across multi-ethnic databases. (A) Hierarchical clustering of Z-score normalized P/LP variant counts for the union of the top 20 genes. Columns are clustered by genetic similarity, revealing two primary clades: a Chinese cluster (ChinaMAP, HuaBiao, WBBC) and a predominantly European (gnomAD, ExAC). (B) Relative allelic enrichment analysis. Counts were normalized by database size to reflect true population frequencies, followed by row-wise Z-score normalization. Annotated color bars indicate inheritance modes: AR (green) and XLR (yellow).

Rank analysis using a bump chart (Supplementary Figure S2) confirmed that *ABCA4* and *USH2A* maintained top-tier rankings across all databases. However, notable ethnic divergences emerged: *RPGR* ranked third in gnomAD exome V4.0.0 but did not appear in the top 10 of any Chinese database. Conversely, *CYP4V2* consistently ranked within the top 10 across ChinaMAP (rank 10), HuaBiao (rank 6), and WBBC (rank 8), yet ranked 91st in gnomAD exomes. Despite variations in cohort size and sequencing depth, the top 10 gene rankings among the three Chinese cohorts showed high consistency, with *CNGA3* and *RDH12* also consistently present in the upper tiers.

### 3.2 Divergent Genetic Architectures of AR and AD-IRDs in the Chinese Cohort

The mutational spectra of IRDs diverged markedly based on inheritance patterns. AR-IRDs were characterized by a highly concentrated mutational landscape, where a small subset of genes accounted for the majority of the pathogenic burden. Notably, the two leading genes, *ABCA4* (n=67) and *USH2A* (n=55), demonstrated extensive allelic heterogeneity, collectively harboring a large proportion of distinct P/LP variants (Table 2). This pattern was consistent across all analyzed Chinese databases.

**Table 2.** Comparative profile of the top 10 AR-IRD-associated genes harboring P/LP variants.

| Top10 Gene | ChinaMAP | HuaBiao | WBBC | NARD | ExAC | gnomAD_exome V2.1.1 | gnomAD_genome V2.1.1 | gnomAD_exome V4.0.0 | gnomAD_genome V4.0.0 |
| --- | --- | --- | --- | --- | --- | --- | --- | --- | --- |
| 1 | *ABCA4*(67) | *ABCA4*(38) | *USH2A*(34) | *ABCA4*(14) | *ABCA4*(277) | *USH2A*(407) | *ABCA4*(127) | *USH2A*(641) | *ABCA4*(296) |
| 2 | *USH2A*(55) | *USH2A*(31) | *ABCA4*(30) | *USH2A*(6) | *USH2A*(258) | *ABCA4*(388) | *USH2A*(96) | *ABCA4*(570) | *USH2A*(265) |
| 3 | *CRB1*(22) | *EYS*(19) | *CRB1*(15) | *EYS*(5) | *CEP290*(123) | *CEP290*(186) | *EYS*(53) | *CEP290*(398) | *EYS*(150) |
| 4 | *EYS*(22) | *ABCC6*(15) | *EYS*(14) | *PDE6B*(4) | *MYO7A*(100) | *MYO7A*(182) | *MYO7A*(51) | *EYS*(344) | *MYO7A*(135) |
| 5 | *CNGA3*(21) | *MYO7A*(14) | *CNGA3*(13) | *ABCC6*(3) | *ADGRV1*(84) | *EYS*(159) | *CEP290*(44) | *MYO7A*(325) | *CEP290*(134) |
| 6 | *CEP290*(19) | *CYP4V2*(12) | *CEP290*(11) | *BEST1*(3) | *ALMS1*(79) | *ADGRV1*(146) | *ABCC6*(42) | *ALMS1*(265) | *ALMS1*(87) |
| 7 | *MYO7A*(18) | *RDH12*(11) | *MYO7A*(11) | *CC2D2A*(3) | *CRB1*(78) | *ALMS1*(135) | *CRB1*(32) | *ADGRV1*(243) | *ABCC6*(84) |
| 8 | *DYNC2H1*(15) | *BEST1*(10) | *CYP4V2*(10) | *CNGA3*(3) | *ABCC6*(71) | *CRB1*(130) | *ADGRV1*(30) | *CDH23*(218) | *ADGRV1*(83) |
| 9 | *CC2D2A*(14) | *CC2D2A*(10) | *RDH12*(10) | *NMNAT1*(3) | *CNGA3*(71) | *NBAS*(110) | *CNGA3*(28) | *CRB1*(199) | *CRB1*(80) |
| 10 | *CYP4V2*(12) | *CEP290*(9) | *ABCC6*(9) | *RDH5*(3) | *NBAS*(66) | *CDH23*(101) | *RPE65*(28) | *PEX1*(157) | *CC2D2A*(78) |

In contrast, AD-IRDs displayed a diffuse genetic distribution with pronounced locus heterogeneity. Most AD genes harbored only a small number of distinct P/LP variants (Table 3). For example, the top-ranked AD genes in ChinaMAP, such as *COL2A1* and *MFN2*, each contained only three distinct variants. Cross-database concordance of top-ranked AD genes was low, contrasting with the consistent dominance observed for AR-IRDs.

**Table 3.**
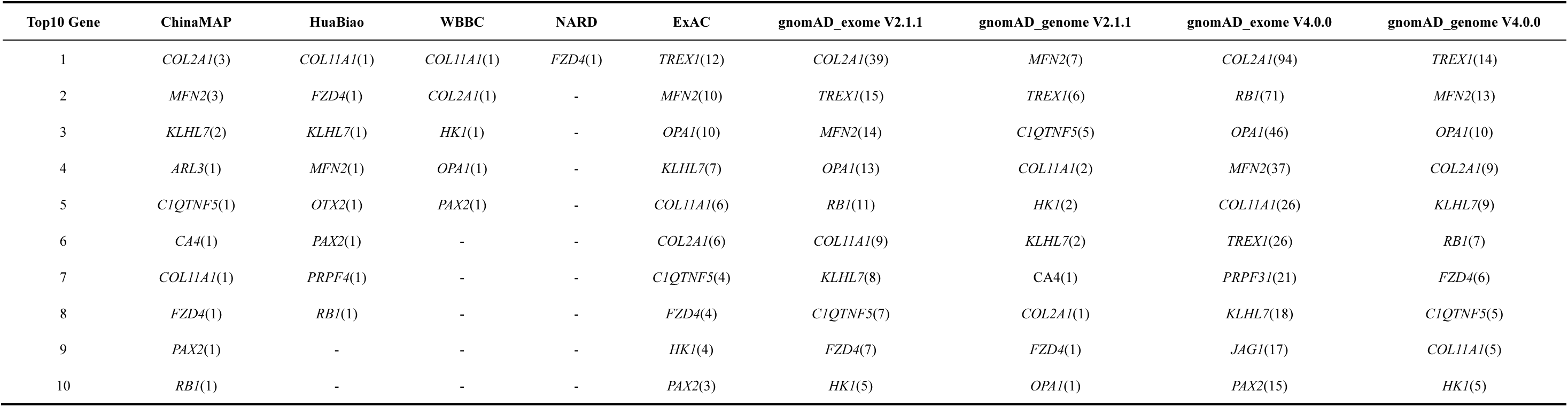
Comparative profile of the top 10 AD-IRD-associated genes harboring P/LP variants.

To mathematically quantify this distributional inequality, we performed Lorenz curve analysis and calculated Gini coefficients (Figure 3). In the ChinaMAP cohort, AR-IRDs showed a deviation from the line of equality (G = 0.501), a pattern also observed in global datasets (G ≈ 0.5), suggesting that the concentrated burden in AR-IRDs is a consistent genomic feature. In contrast, AD-IRDs in ChinaMAP displayed a more dispersed architecture (G = 0.247), lower than aggregated global values, reflecting the high locus heterogeneity in the Chinese population. Notably, in global databases, AD-IRDs paradoxically showed higher concentration (G ≈ 0.6) than AR-IRDs (G ≈ 0.5), likely due to the presence of high-frequency founder mutations in a few AD genes (e.g., RHO, PRPF31) within European-dominant cohorts. Consistently, inter-database correlation coefficients for AR genes were higher than for AD genes (Supplementary Figure S3), indicating that the mutational spectrum of AR-IRDs is more reproducible across independent cohorts.

**Figure 3.**
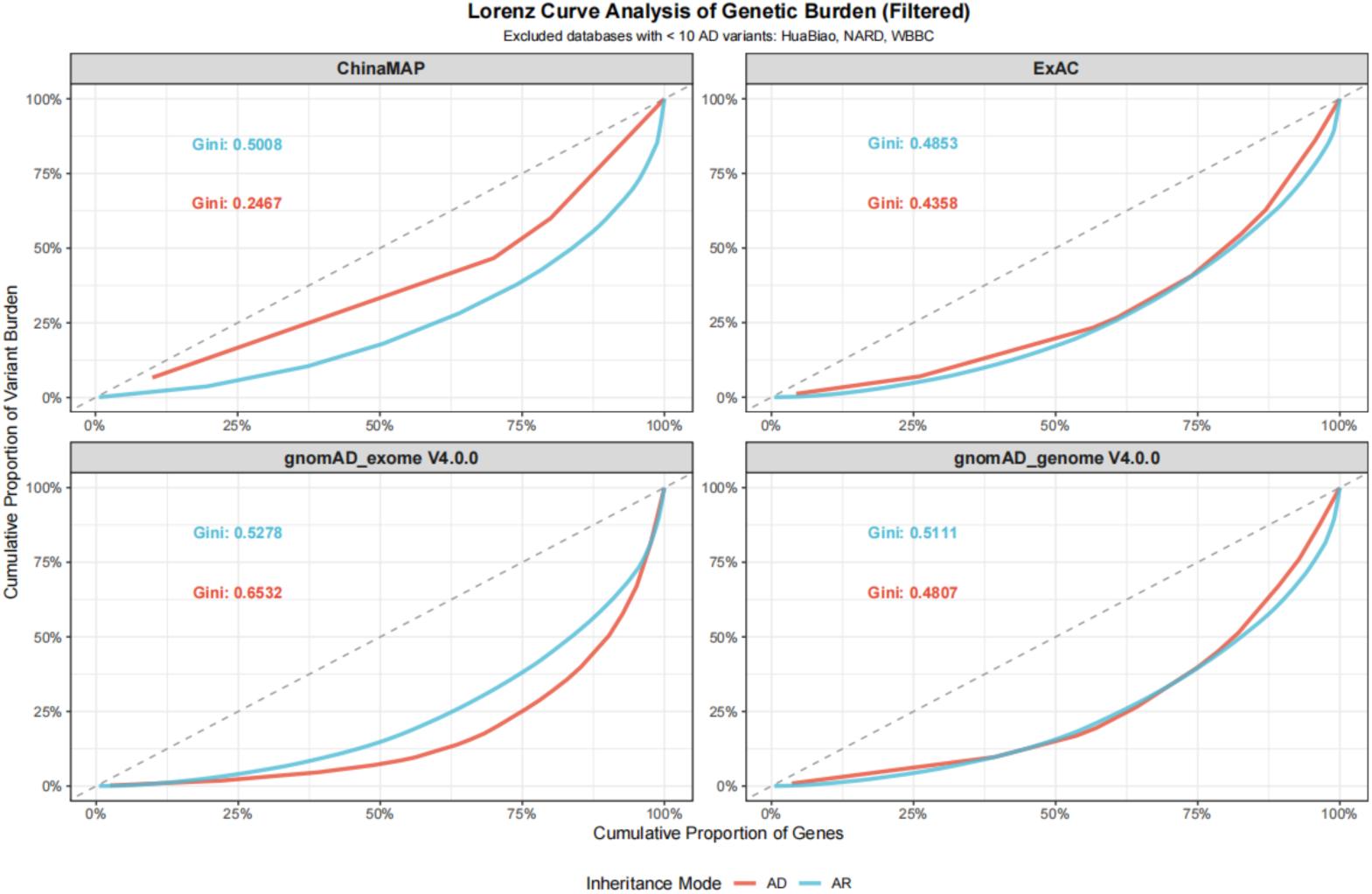
Lorenz Curve Analysis of Distribution of Distinct P/LP Variant Types in AR and AD-IRD Genes. The X-axis represents the cumulative proportion of genes (ranked by increasing number of distinct P/LP variants); the Y-axis represents the cumulative proportion of distinct variants. The dashed diagonal line indicates perfect equality (Gini = 0). AR-IRDs (blue) show a larger deviation (Gini = 0.501 in ChinaMAP), indicating concentration; AD-IRDs (red) lie closer to the diagonal (Gini = 0.247), indicating locus heterogeneity. Databases with insufficient AD variants (total distinct variants < 10) were excluded.

We further assessed the stability of allelic diversity across seven databases by calculating the coefficient of variation (CV) for the normalized count of distinct variants per gene (Figure 4). AR genes demonstrated relatively high consistency in their mutational spectra across databases (median CV ≈ 0.77), whereas AD genes exhibited significantly higher volatility (median CV ≈ 1.72; P < 0.0001, Mann-Whitney U test). Permutation tests confirmed the robustness of these findings (Supplementary Figure S5).

**Figure 4.**
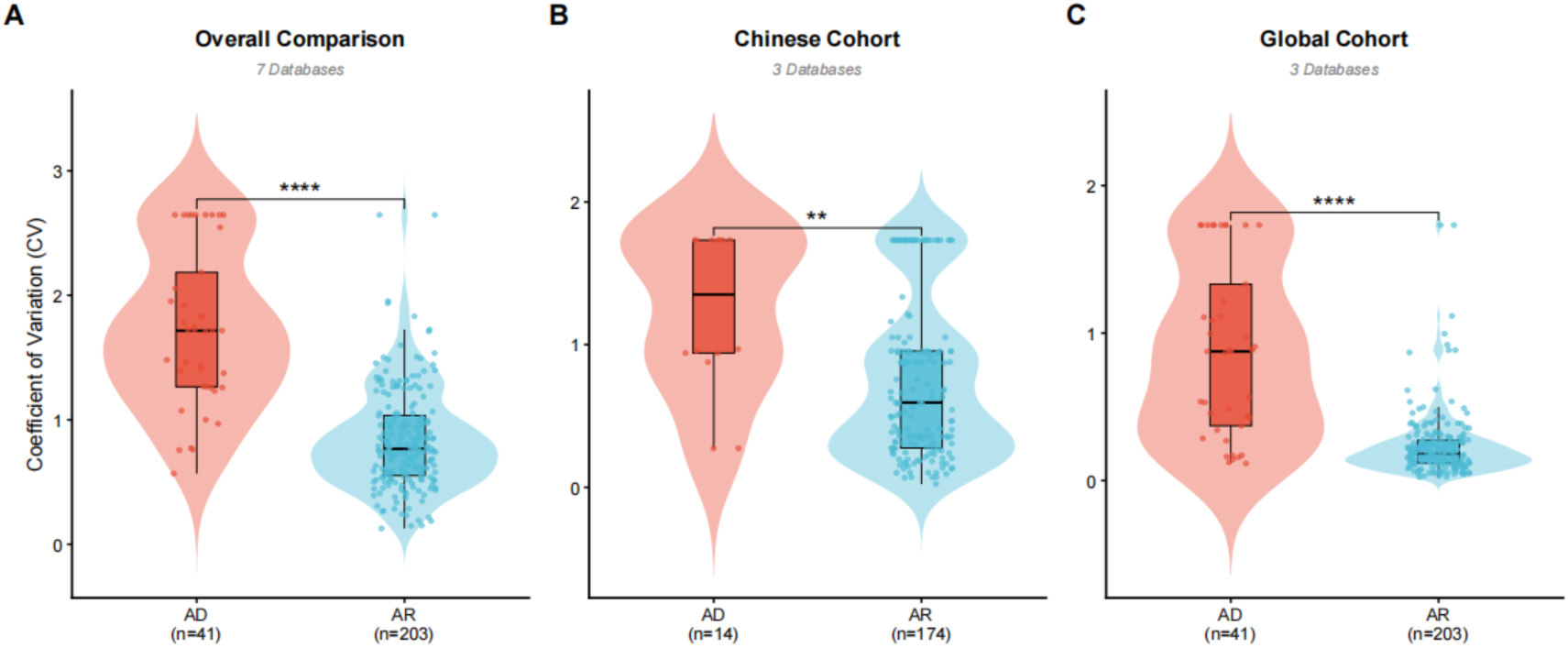
Cross-Cohort Validation of Allelic Diversity Stability. Violin plots comparing the coefficient of variation (CV) of distinct P/LP variant counts between AR (n = 203) and AD (n = 41) genes across (A) all seven databases, (B) Chinese cohorts only, and (C) global cohorts only. AD genes consistently show higher variability than AR genes (P < 0.01, Mann-Whitney U test).

### 3.3 Population-Specific Genetic Variations Revealed by the Allele Frequency Spectrum

Analysis of variant frequencies in the ChinaMAP cohort revealed a distinct set of high-prevalence P/LP variants (Table 4). The most prevalent was the missense substitution in *CNGB1*, c.C1589G (p.P530R), which exhibited an AF of 0.006139 in ChinaMAP and was consistently validated across independent Chinese cohorts (HuaBiao: 0.004840; WBBC: 0.006250).

**Table 4.**
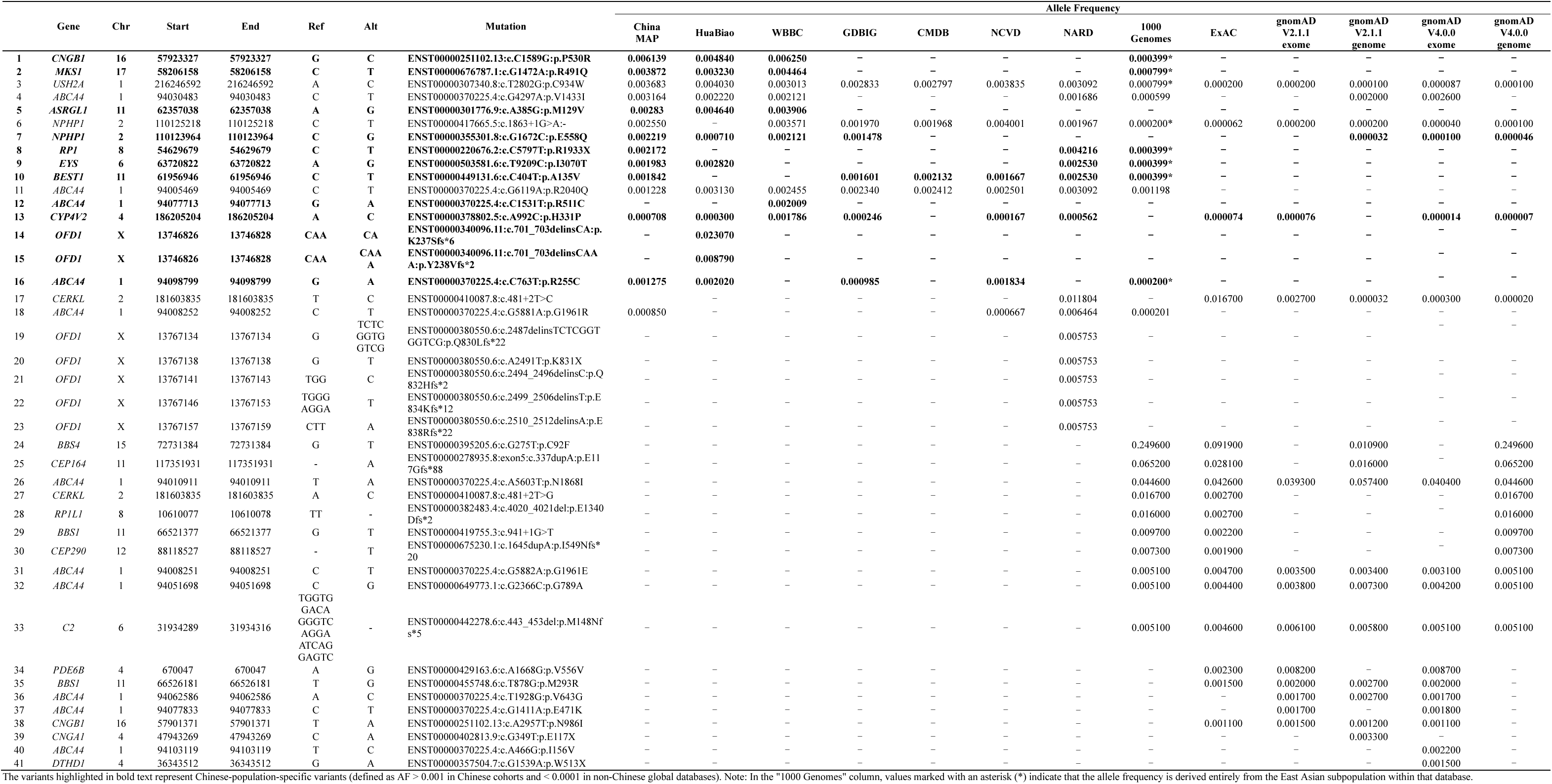
Comparison of the top 10 IRD-associated genes ranked by allele frequency across major population databases.

Applying our specific criteria for population specificity (AF > 0.001 in Chinese databases and < 0.0001 in non-Chinese global databases), we identified a subset of variants representing distinct local genetic features (shown in bold in Table 4). In addition to *CNGB1*(p.P530R), this group includes *MKS1*(p.R491Q), *ASRGL1*(p.M129V), *NPHP1*(p.E558Q), *RP1*(p.R1933X), *EYS*(p.I3070T), *BEST1*(p.A135V), *ABCA4*(p.R511C), *CYP4V2*(p.H331P), *OFD1*(p.K237fs*6 and p.Y238Vfs*2) and *ABCA4*(p.R255C). Collectively, these variants, particularly *MKS1* (p.R491Q) and the *NPHP1* mutations, demonstrated robust frequencies within China while being virtually undetectable in broad global datasets like gnomAD, confirming their status as population-specific genetic markers. The pronounced enrichment of these top mutations in Chinese cohorts is clearly illustrated in Figure 5A (heatmap) and Figure 5B (allele frequency comparison). For instance, *CNGB1*(p.P530R) and *MKS1*(p.R491Q) were absent in gnomAD, suggesting a potential founder effect unique to Chinese or broader East Asian ancestry. Several other top-ranked variants, including *NPHP1*(p.E558Q) and those in *RP1* and *EYS*, were exclusively detected within the gnomAD East Asian subpopulation, further demonstrating ancestral enrichment.

**Figure 5.**
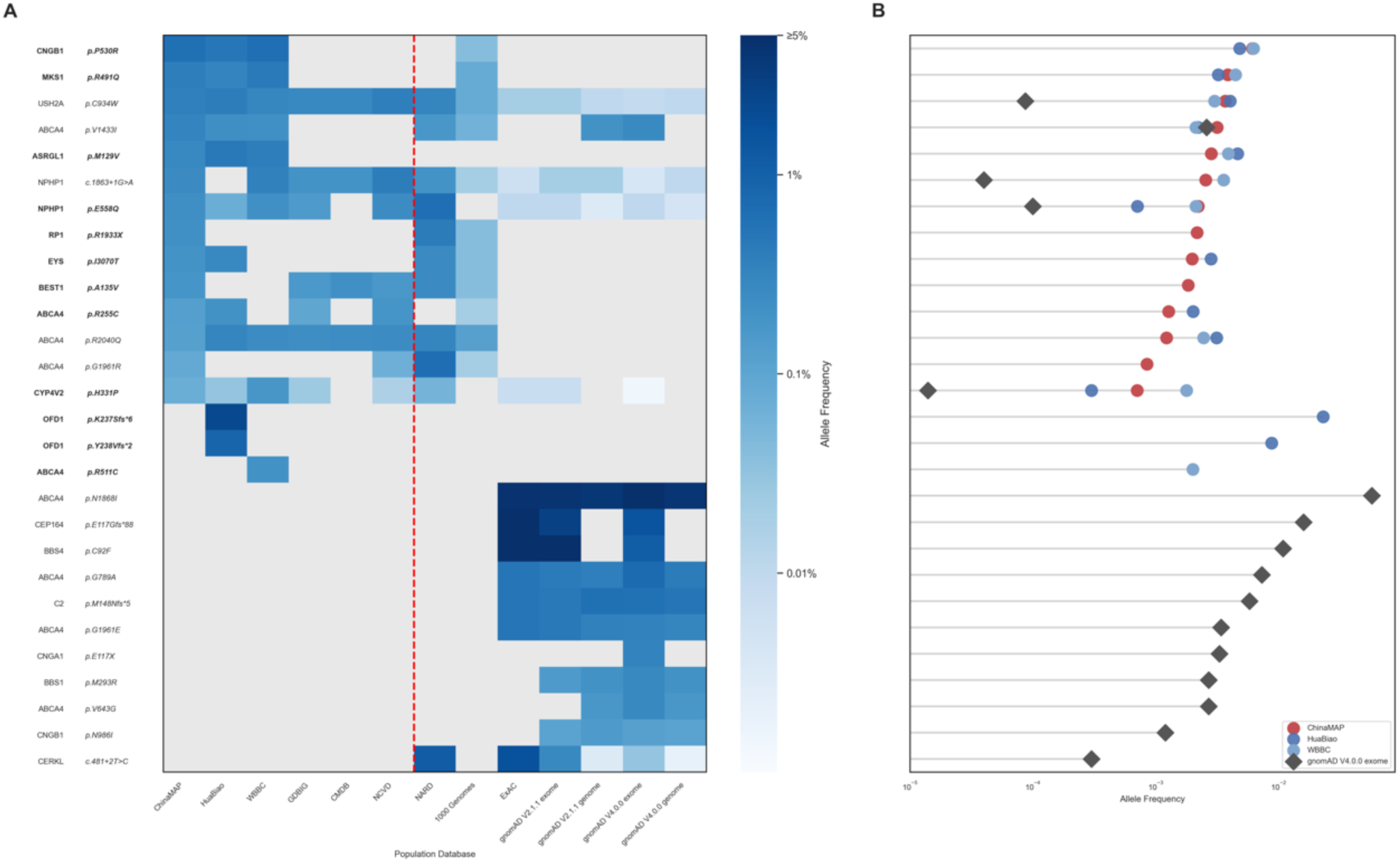
Comparison of Allele Frequencies for IRD Variants Across Major Population Databases. (A) Heatmap visualization of the allele frequency spectrum for the top pathogenic/likely pathogenic variants. Frequencies are compared across multiple Chinese cohorts (ChinaMAP, HuaBiao, WBBC, etc.) and predominantly European databases (gnomAD, ExAC, 1000 Genomes). The color intensity reflects the relative allele frequency. (B) A plot illustrating the allele frequencies of these high-frequency variants. The allele frequencies in Chinese databases (ChinaMAP, HuaBiao, and WBBC) are contrasted with the global gnomAD V4.0.0 exome data.

### 3.4 Refined Estimates of CF and GP Analyses in the Chinese Population

To quantify the overall genetic burden of AR-IRDs in the Chinese population, we calculated the cumulative CF and total GP using the ChinaMAP and WBBC cohorts. The cumulative CF for the 209 AR-IRD genes was 1 in 5.60 in ChinaMAP (Supplementary Table 3) and 1 in 5.68 in WBBC (Supplementary Table 4), demonstrating high cross-cohort stability. Based on these CFs, the theoretical total GP was estimated at 1 in 2,624.67 for ChinaMAP and 1 in 2,612.86 for WBBC. These results provide a population-specific reference for AR-IRD burden in the Chinese population, underscoring the importance of using ancestry-matched data for accurate risk assessment.

## 4. Discussion

This study systematically evaluated the pathogenic variant spectrum of IRDs in the Chinese population through an integrated analysis of variants from ChinaMAP and multiple domestic and international public databases. The findings not only validate and expand upon previous discoveries from clinical cohorts but also provide a genetic reference dataset for developing precise diagnostic strategies, genetic counseling protocols, and future therapeutic pathways tailored to the Chinese population.

A key finding of our investigation is that the pathogenic variants in AR-IRDs are concentrated in a small subset of genes. The genes *ABCA4* and *USH2A* were identified as the principal contributors to the genetic burden, known to be the predominant causes of Stargardt disease^[21]^ and retinitis pigmentosa^[22]^, respectively. Although the rank of most commonly reported genes showed minor variations compared to a study of 1,229 Chinese patients with IRDs^[23]^, our observation was robustly validated across multiple independent Chinese cohorts and aligns with previous, smaller-scale clinical studies in China^[13,24]^. However, the strength of our study lies in its use of large-scale variant data from the general population, which provides population-level evidence that more accurately reflects the true distribution and burden of these mutations, thereby establishing a critical basis for public health assessments. The dominance of these high-frequency pathogenic genes contrasts with patterns observed in European descent-centric databases like gnomAD, where other genes, such as *RPGR*, hold higher ranks. Conversely, genes such as *CYP4V2* and *RDH12* exhibited distinct population specificity. *CYP4V2* consistently ranked within the Top 10 across all Chinese datasets but was notably absent from the upper tiers of global databases. This divergence highlights the geographic and ancestral stratification of IRD genetics and underscores the critical need to move beyond reliance on global reference databases, which can lead to diagnostic inaccuracies and an under-appreciation of key pathogenic drivers in specific ethnic groups^[10]^, as hypothesized in our introduction.

Our analysis mathematically quantified the dichotomy between AR- and AD-IRDs. The high Gini coefficient (G ≈ 0.5) for AR-IRDs supports the efficiency of tiered diagnostic screening, where a panel targeting a few high-frequency genes can achieve a high theoretical detection rate, in line with previous findings^[25]^. In contrast, AD-IRDs exhibit high locus heterogeneity (G = 0.2467), reflecting a highly dispersed mutational spectrum where the pathogenic load is distributed across numerous genes, each typically harboring only a few variants. This is consistent with global findings that AD-IRDs involve dozens of genes, most with low mutation frequencies^[5]^. This distribution mandates a shift in diagnostic triage: suspected AD-IRDs require comprehensive whole-exome sequencing (WES) or WGS to capture the sparse mutational spectrum, as targeted panels are likely to suffer from low sensitivity in this context. Furthermore, our findings caution against directly applying panels designed for other ethnicities, as key genes in global databases, like *TREX1*, are not prominent in the Chinese cohort, highlighting the critical need for population-specific diagnostic references^[26]^. Permutation tests further confirmed that the stability of the AR-IRD mutational spectrum is a robust biological feature, whereas the high volatility observed for AD-IRDs reflects both stochastic mutation occurrence and population-specific divergence.

Notably, the presence of such variants is not confined to clinical cohorts; our study identified them within a general population. This reflects the latent pathogenic load of IRDs, which can be attributed to variable expressivity or presymptomatic stages in late-onset conditions. Therefore, our calculated genetic prevalence provides an upper bound of the disease burden. The discrepancy between genetic predictions and clinical observation may point to the existence of protective genetic modifiers or incomplete penetrance, which warrants future long-term natural history studies.

Characterizing the distinct genetic spectrum of IRDs in the Chinese population is a prerequisite for regionalized precision medicine. A prime example is the *CYP4V2* gene, closely associated with Bietti’s crystalline dystrophy, which shows a significantly higher pathogenic burden in East Asian populations^[27,28]^. Our study provides baseline data for this gene, documenting a high variant allele frequency in the general Chinese population (e.g., 0.000708 in ChinaMAP). This study also identified several other pathogenic variants significantly enriched in the Chinese population. For instance, the CF of the *ABCA4*(p.R255C) variants is much higher than in other global populations. Although previous studies suggested the presence of this mutations in small cohorts of Chinese Stargardt disease patients^[29]^, our work provides large-scale, population-based evidence confirming its high carrier rates and offering compelling evidence of founder effects for *ABCA4*-related retinopathies in this population. Furthermore, we identified a distinct set of variants including *MKS1*, *NPHP1*, and *RP1* that are uniquely prevalent in China but absent or extremely rare in non-East Asian global databases.

Of particular interest is the *CNGB1*(p.P530R) variant, which showed a high allele frequency (∼0.006) in Chinese cohorts and was virtually absent elsewhere. Although we did not perform functional assays, its localization to a conserved cyclic nucleotide-binding domain suggests a potential impact on protein function, warranting further experimental investigation. Additionally, our exploratory analysis hinted at possible regional stratification within China—for example, *CNGB1*(p.P530R) appeared more frequent in Northeast China, while *ASRGL1*(p.M129V) was relatively enriched in Central and Southern China. These observations, though preliminary and requiring validation in larger geographically tagged cohorts, suggest that founder effects may have acted differently across subpopulations, and highlight the potential value of regionally optimized genetic screening strategies in a country as vast and diverse as China. The identification of these population-specific variants exposes the limitations of relying on ethnically mismatched reference controls. Integrating this robust dataset is critical for variant interpretation under ACMG/AMP guidelines^[30–32]^, empowering clinicians to distinguish pathogenic mutations from polymorphisms accurately. Ultimately, these findings serve as an evidence base for designing high-yield, cost-effective genetic screening panels tailored for the Chinese population, thereby facilitating expanded carrier screening, preimplantation genetic testing, and the broader implementation of regionalized reproductive health strategies^[33,34]^.

In addition, our calculation of the cumulative CF (1 in 5.60) and total GP (1 in 2,624.67) of ChinaMAP provides a refined estimate of the public health burden of AR-IRDs in China. When compared to domestic data, our estimates are similar to those reported in the WBBC cohort, though the two cohorts have different sizes: ChinaMAP consists of 10,588 individuals, while the WBBC cohort has 4,480 individuals. This slight discrepancy may stem from regional stratification: for instance, the WBBC cohort, primarily representing Southern Chinese populations, may exhibit a different distributional density of certain founder mutations compared to the nationally-representative ChinaMAP dataset. In global comparison, regarding global comparison, our results differ from the figures reported for East Asians in gnomAD (V2.1.1)^[1]^. It is crucial to acknowledge that the “East Asian” ancestry group in gnomAD is a broad aggregate of diverse populations, including Korean, Japanese, and Chinese individuals, and thus cannot serve as an accurate representation for the specific genetic landscape of the Chinese population. Both sample size and specific population composition fundamentally influence the accuracy of CF and GP calculations. Given that ChinaMAP benefits from a larger scale and a focused Chinese cohort, we conclude that our estimates offer a more credible and representative reference for the Chinese population. These findings provide a critical evidence base for healthcare authorities to implement targeted population-wide carrier screening, potentially reducing the socioeconomic burden of IRDs.

The primary strengths of this study lie in its large sample size and the use of variants from high-depth WGS data from the ChinaMAP cohort. By cross-validating variants against multiple databases, this research provides a high-fidelity, population-level map of the IRD genetic landscape in the general population compared to previous studies based on smaller, clinic-based cohorts or exome sequencing. Nevertheless, several limitations should be acknowledged. First, from a statistical and discovery perspective, the “zero-inflation” observed in our AD-IRD analysis indicates that even with a cohort of N=10,588, we are only beginning to saturate the rare variant discovery curve. The stochastic nature of dominant mutations suggests that a significant portion of the “missing heritability” remains to be uncovered. Future studies with even larger cohorts and the integration of long-read sequencing will be essential to resolve complex genomic regions and identify variants beyond the reach of standard short-read WGS.

Second, regarding variant spectrum and cohort composition, our reliance on a general population cohort, while excellent for establishing a baseline, may inevitably omit ultra-rare or private mutations that manifest exclusively within specific IRD pedigrees. Furthermore, although ChinaMAP is geographically expansive, it may not fully capture the pan-ethnic diversity of China’s 56 ethnic groups, particularly those in isolated geographical sub-regions or minority clusters with distinct genetic backgrounds.

Third, concerning mutational complexity, our current pipeline focuses primarily on single-nucleotide variants and small insertions-deletions (indels). Due to the inherent challenges in calling large-scale events from short-read data without extensive raw sequence re-processing, we likely underestimated the contribution of structural variations and copy number variants, which are increasingly recognized as pivotal drivers of IRD pathogenesis.

Finally, looking toward clinical translation, while our pathogenicity assessments strictly adhere to ACMG/AMP guidelines, large-scale functional experimental validation remains the gold standard to definitively confirm these in silico predictions. Future research should prioritize the integration of this population-level genomic data with deep clinical phenotyping to refine our understanding of genotype-phenotype correlations and incomplete penetrance. Ultimately, combining these ancestry-specific genetic maps with pharmacogenomic profiles will provide a critical roadmap for the development of gene-based therapies, advancing the frontier of precision ophthalmology in China.

## 5. Conclusion

This study delineates the comprehensive genetic landscape of IRDs in the Chinese population by leveraging large-scale whole-genome sequencing data. It reveals a clear architectural dichotomy between inheritance patterns: AR-IRDs are characterized by a predominance of recurrent pathogenic alleles, whereas AD-IRDs exhibit pronounced locus heterogeneity with a dispersed mutational spectrum. By identifying Chinese-specific pathogenic variants and providing refined estimates of carrier frequency and genetic prevalence, this work establishes a robust, population-specific reference for the genetic epidemiology of IRDs. These findings serve as an evidence base for optimizing diagnostic strategies and empowering genetic counseling, thereby guiding public health initiatives, including carrier screening programs, to mitigate the disease burden of IRDs in China.

## Data Availability

http://www.mbiobank.com/

https://wbbc.westlake.edu.cn/downloads.html

https://www.biosino.org/wepd/download

http://bigdata.ibp.ac.cn/NyuWa_variants/search.php

http://gdbig.bigcs.com.cn/

https://cmdb.bgi.com/

https://gnomad.broadinstitute.org/downloads

https://www.internationalgenome.org/

https://nard.macrogen.com/

## Acknowledgements

The authors thank the following databases and their respective teams for sharing data in various forms: ChinaMAP, gnomAD, 1000 Genomes, ExAC, WBBC, HuaBiao, NCVD, GDBIG, CMDB, and NARD. We also gratefully acknowledge all other data contributors and individuals who assisted with this study.

## Author contributions

BZ conceived the original idea and designed the experiments. BZ performed the data analyses, including variant pathogenicity assessment, carrier frequency and genetic prevalence estimation. SZ performed the statistical analyses and cross-database comparisons that supported the main figures. ZC analyzed the results, interpreted the findings, and wrote the original draft of the manuscript. BZ, SZ, and WD reviewed, revised, and edited the manuscript for important intellectual content. WD provided funding acquisition, resources, and overall supervision of the project.

## Data availability

All data are obtained from publicly accessible sources. Should additional data be required, interested parties are encouraged to contact the author; any reasonable requests will be granted.

---

**Supplementary Figure S1.**
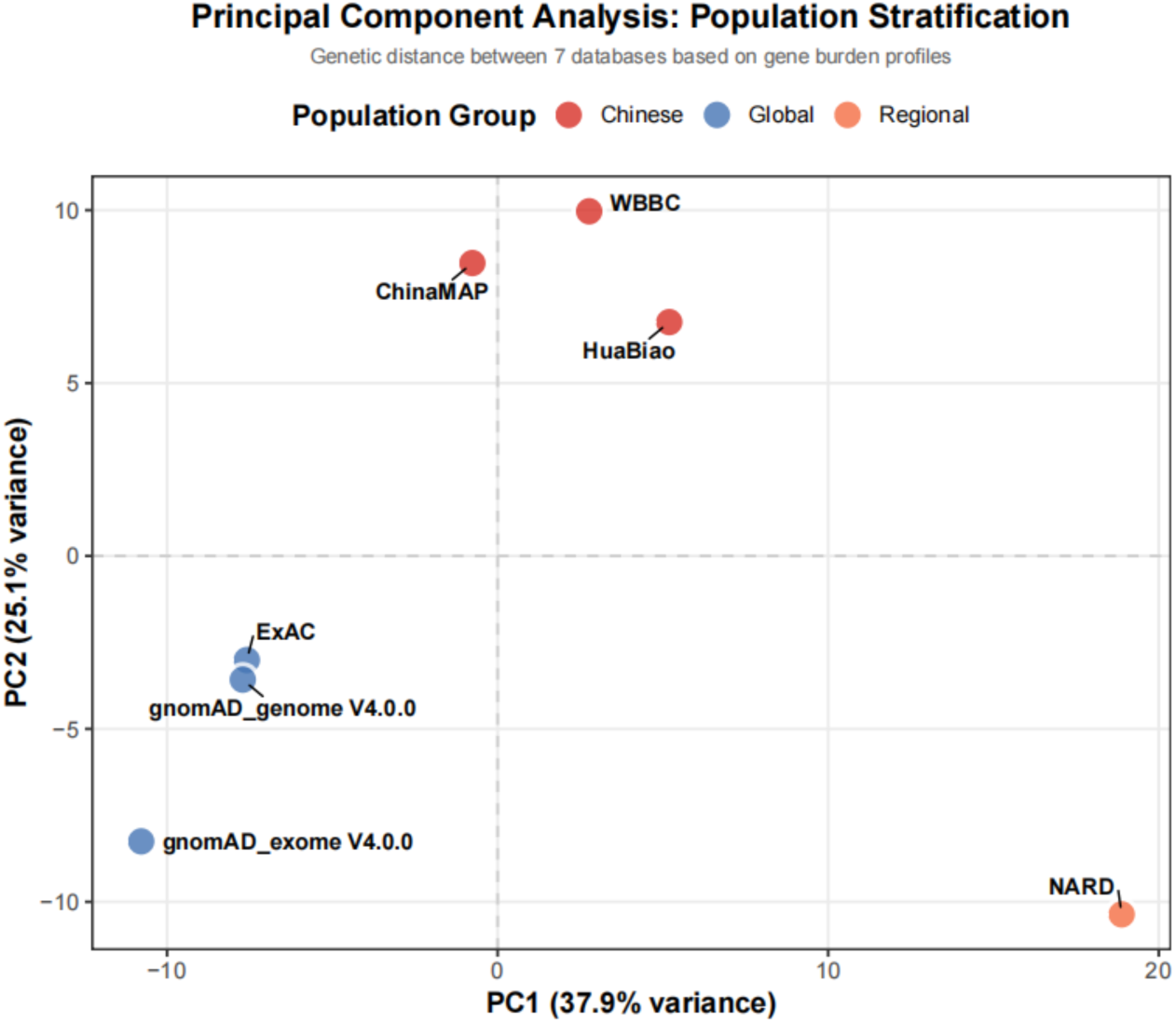
Population-Specific Genetic Stratification of IRD Associated Genes. Principal Component Analysis (PCA) Plot. Databases are projected onto the first two principal components (PCs), which collectively explain 63.0% of the total genetic variance.

**Supplementary Figure S2.**
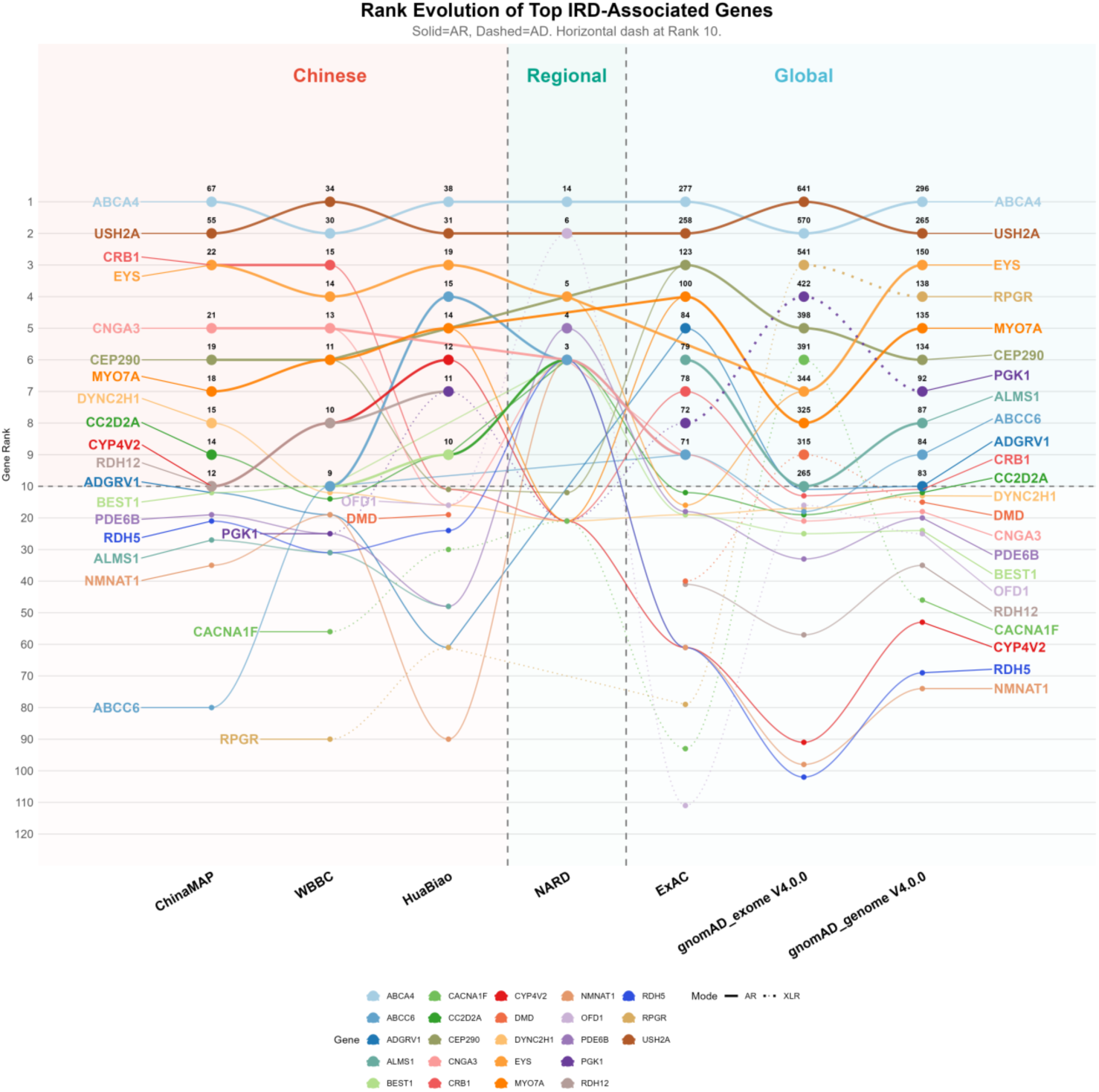
Rank Evolution of Top IRD-Associated Genes. Genes are ranked by the total number of distinct P/LP variants per database. The x-axis shows databases grouped into Chinese (ChinaMAP, WBBC, HuaBiao), regional (NARD), and global (gnomAD, ExAC) cohorts. The y-axis uses a compressed scale for ranks >10 to highlight top-tier genes. Bold curves indicate genes consistently ranked within the top 10; thinner curves represent lower rankings. The horizontal dashed line marks the top-10 threshold. Discontinuous lines indicate missing detection in a given database.

**Supplementary Figure S3.**
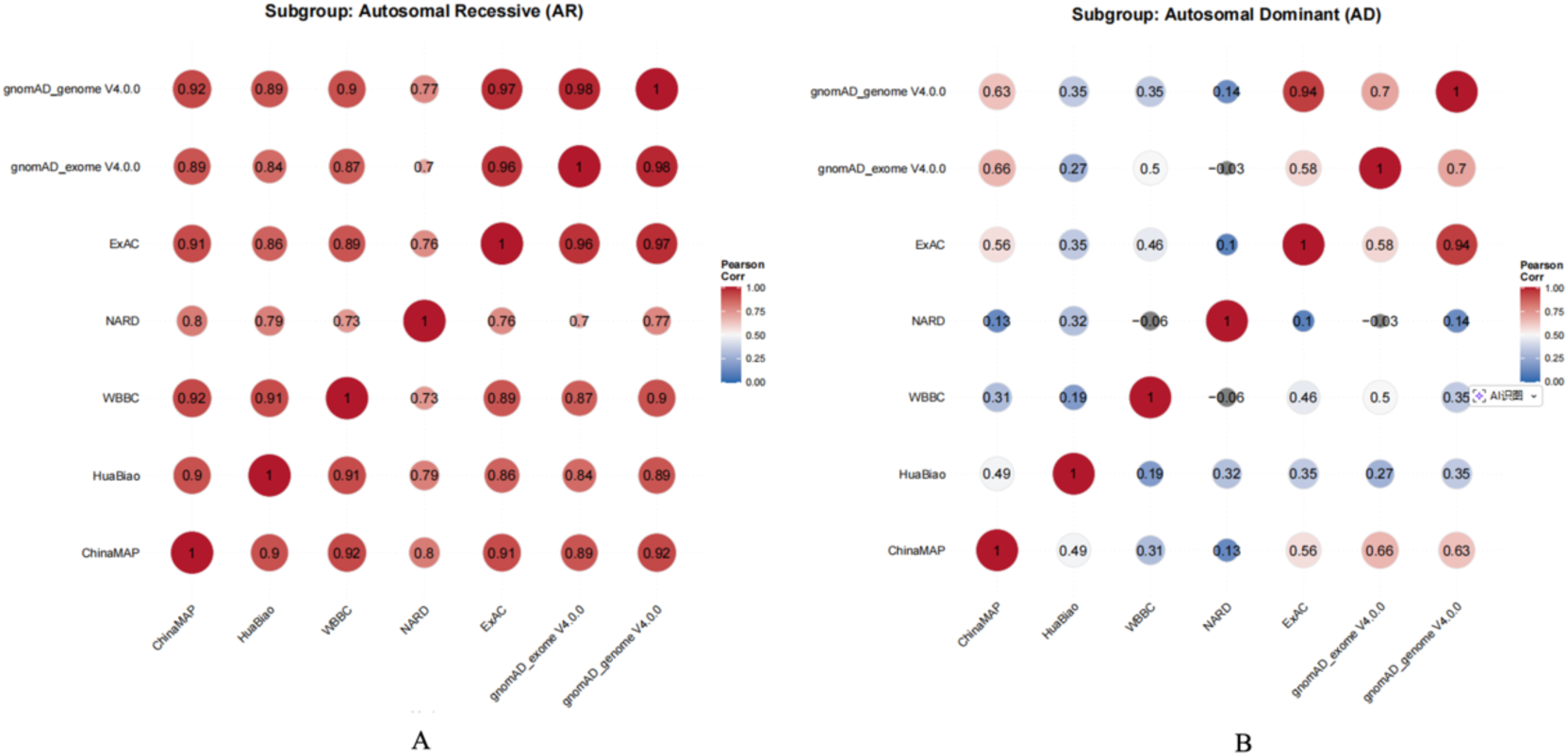
Inter-Database Correlation Analysis of Pathogenic Variant Distributions. The correlation heatmap illustrates the consistency of genetic landscapes across multiple cohorts, stratified by inheritance patterns. (A) AR genes Matrix, (B) AD genes Matrix. Spearman correlation coefficients were calculated based on the relative allelic diversity. Hierarchical clustering of the matrices reveals a primary division based on population origins.

**Supplementary Figure S4.**
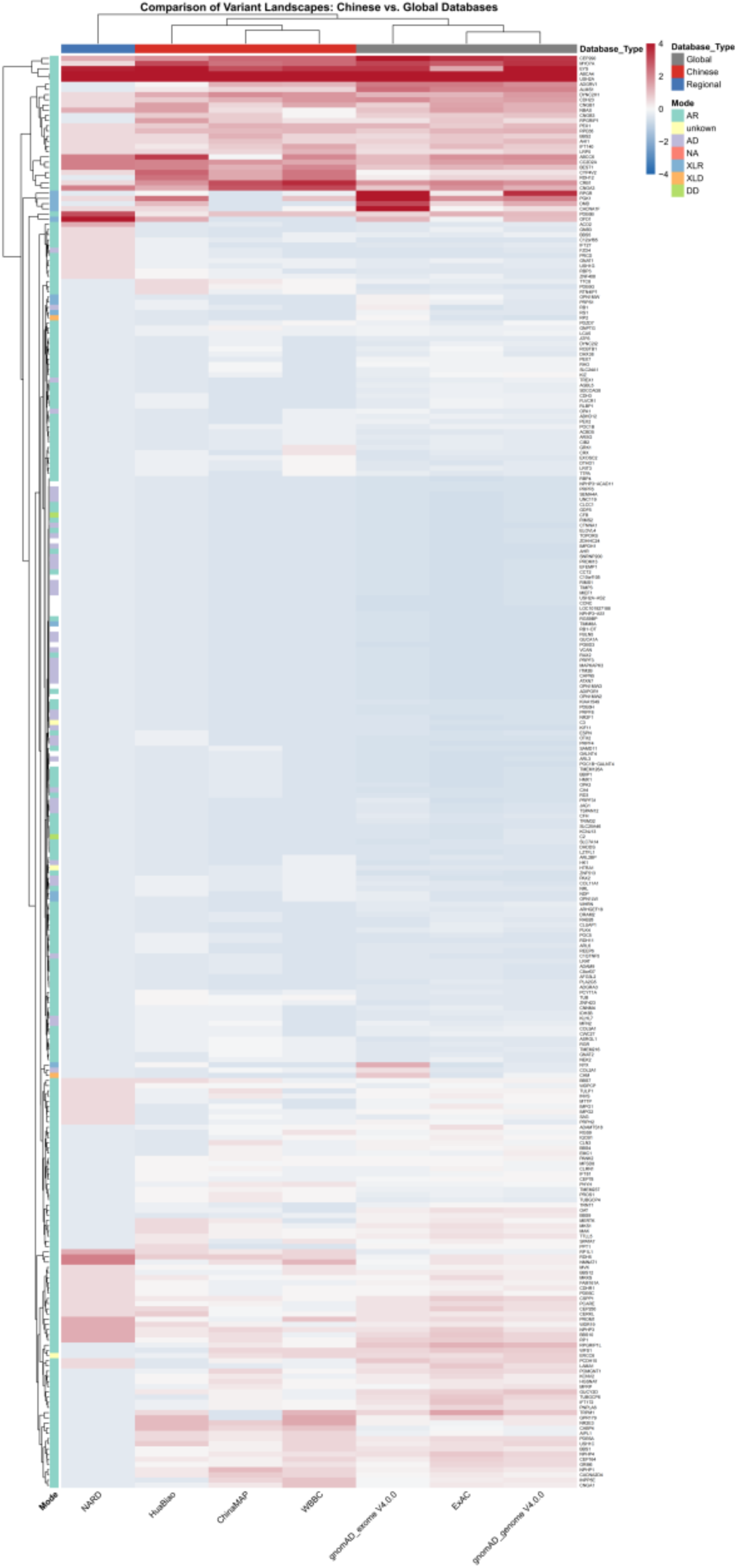
Hierarchical clustering of normalized variant counts separates Chinese cohorts (ChinaMAP, HuaBiao, WBBC) from predominantly European cohorts (gnomAD, ExAC). Color bars indicate inheritance modes.

**Supplementary Figure S5.**
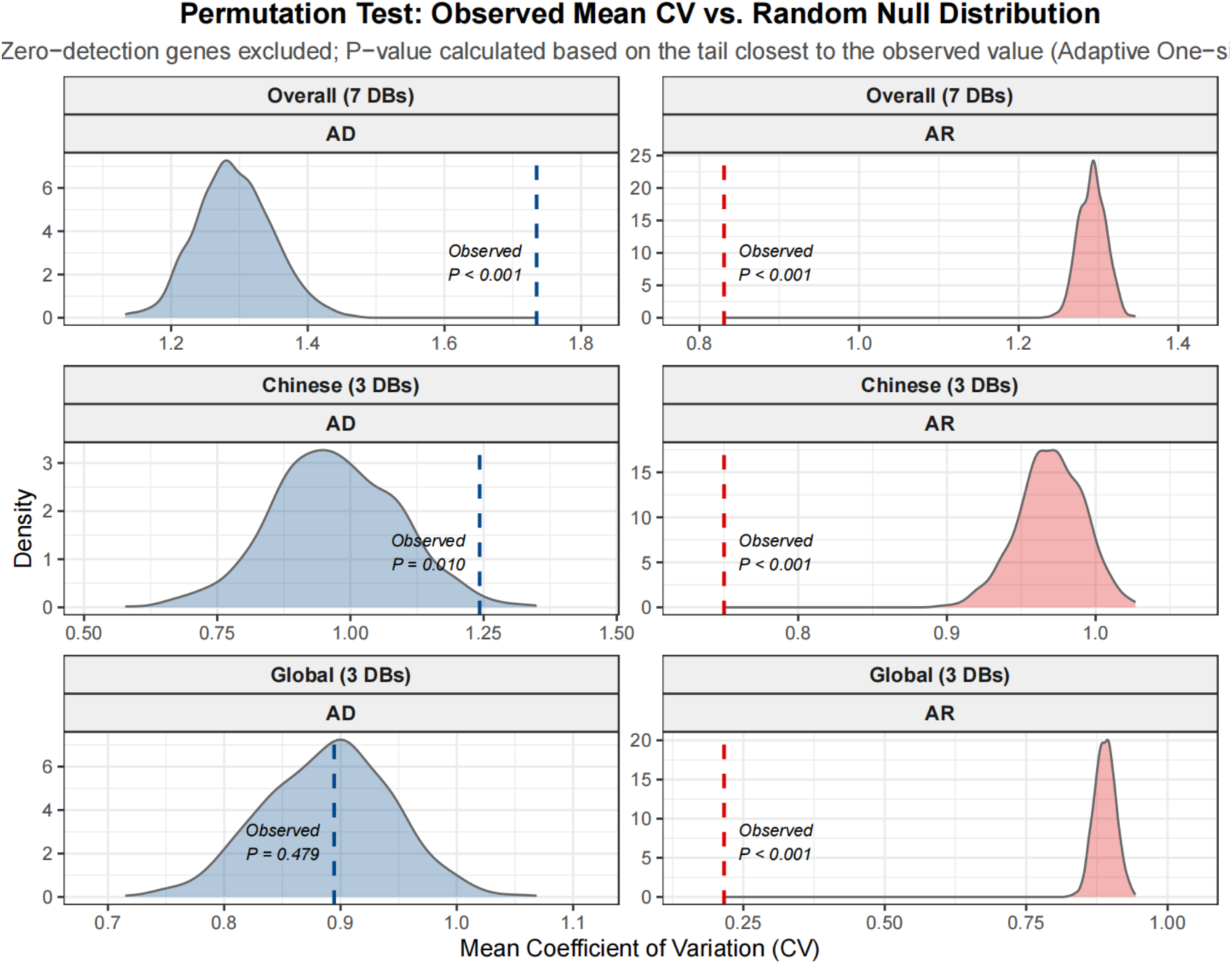
Permutation-Based Validation of Architectural Constraints and Data Sparsity. Density plots show null distributions (grey) from 1,000 random permutations; dashed vertical lines indicate observed mean CVs. Data are stratified by cohort (Overall, Chinese, Global) and inheritance mode (red: AR; blue: AD). P-values are empirical one-sided probabilities. AR genes show consistently lower CV than null (P < 0.001). AD genes show context-dependent patterns: hyper-volatility in Overall and Chinese cohorts, but no significant deviation in Global cohorts

